# Cross-binding antibodies capable of neutralizing diverse orthohantaviruses are produced in response to Puumala virus infection

**DOI:** 10.1101/2025.03.04.25323331

**Authors:** Jordan Clark, Stefan Hatzl, Kirill Vasilev, Robert Andreata-Santos, Jeremy S. Yong, Eva Mittler, Ezgi Kasikci, Kartik Chandran, Viviana Simon, Robert Krause, Florian Krammer

## Abstract

**Background:** Orthohantaviruses (hantaviruses) are emerging rodent-borne pathogens that can cause severe human disease. They are present on multiple continents and are responsible for thousands of human cases per year, predominantly in China. Despite this, no vaccines or licensed therapeutics are available, and the immunological response to infection is poorly characterized. This study aimed to analyze the humoral response to PUUV infection to inform future studies focusing on the production of therapeutic monoclonal antibodies and vaccination strategies.

**Methods:** Serum was obtained from a cohort of 23 patients hospitalized with Puumala virus (PUUV) infection at four time points, covering the early acute, late acute, early convalescent, and late convalescent stages of the disease. The humoral responses at each time point were quantified, and cross-binding, cross-neutralizing antibody responses were investigated. Serum cytokine levels were also interrogated, and expression was correlated with humoral outputs.

**Findings:** PUUV infection elicited a robust anti-PUUV neutralizing antibody response. However, cross-reactive antibodies that were capable of binding diverse hantaviruses were also induced in late convalescence. Modulations in the abundance of IgG subclasses were also evident following infection, with significant differences present months after infection.

**Interpretation:** This study demonstrates that broadly reactive anti-hantavirus antibodies are produced in response to Old-World hantavirus infection, but predominantly months after recovery. As this is concomitant with changes in IgG subtypes, our results suggest that PUUV antigen persists in humans post-infection, which promotes prolonged class-switching and somatic hypermutation, favoring conserved epitopes.

## Introduction

Orthohantaviruses (Hantaviruses, family: *Hantaviridae,* order: *Bunyavirales*) are rodent-borne, negative-sense, segmented RNA viruses that are found worldwide. These viruses are associated with a specific reservoir host and can be geographically and phylogenetically divided into New-World and Old-World hantaviruses. Hantaviruses naturally infect rodent, shrew, mole, and bats, resulting in avirulent, presumably lifelong infections. Although these reservoir hosts do not exhibit disease signs, they continually shed the virus in their urine, feces, and saliva. These excreta can then become desiccated and, when disturbed, can be inhaled by humans or directly inoculated into broken skin, mucous membranes, or eyes. When infecting humans, viruses harbored by old-world mice and rat species (*Muridae*), such as Seoul virus (SEOV), Hantaan virus (HTNV), and Dobrava-Belgrade virus (DOBV) can cause hemorrhagic fever with renal syndrome (HFRS). Depending on the hantavirus species responsible, HFRS can result in case fatality rates of 1-10%, and there are 150-200,000 cases per year, predominantly in China^1,2^. New-world hantaviruses such as Andes virus (ANDV) and Sin Nombre virus (SNV) are spread by new-world rats and mice (*Sigmodontinae* & *Neotominae*) which are found in the Americas. New-world hantaviruses can cause hantavirus cardiopulmonary syndrome (HCPS), a severe disease that has case fatality rates ranging from 30-60%. Conversely, infections with hantaviruses that are found in voles, lemmings and muskrats (*Arvicolinae*) such as Puumala virus (PUUV) can cause a milder form of HFRS known as *nephropathia epidemica* (NE), which has a lower case fatality rate of ∼0.1%^3,4^. PUUV is the most common cause of hantavirus infection in Europe and was responsible for 98% of laboratory-confirmed infections in the European Union in 2020. Of the European countries, Finland and Germany exhibit the highest case numbers, accounting for 45% and 29% of the European cases between 2016 and 2020 respectively, while other central European countries such as Austria, Slovenia, and Slovakia account for approximately 0.1%-12.1% of European cases^5^. PUUV cases in Europe fluctuate and depend on bank vole populations, which are, in turn, influenced by complex environmental factors such as climate variation and predator populations^5–11^. While PUUV is associated with low case fatality rates, infection can result in debilitating symptoms such as increased vascular permeability, acute kidney injury, and thrombocytopenia which often necessitate hospitalization and potentially intensive care unit admission^12–16^. Although most patients recover without long-term sequelae, rare cases of hypopituitarism have been reported^17^. Several biomarkers have been investigated in patients suffering from PUUV and include: interleukin (IL)-6, C-reactive protein (CRP), pentraxin-3 (PTX3), indoleamine 2,3-dioxygenase (IDO), soluble urokinase-type plasminogen activator (uPAR), and many others^18,19^. While PUUV infections are common in Europe, no targeted treatments are available, and no approved vaccines are currently available for the prevention of hantavirus disease outside of China and South Korea. While it is presumed that PUUV infection elicits life-long immunity, the humoral responses to infection are poorly understood. The glycoprotein of PUUV is encoded by the medium (M) genomic segment, which is translated as a single polypeptide. This is post-translationally proteolytically cleaved into two separate proteins, Gn and Gc, which interact to form heterodimers^20^. These heterodimers interact with adjoining dimers to form tetrameric spikes on the virion surface, which mediate cell entry^21^, and are thus both targets of neutralizing antibodies. Recent therapeutic intervention strategies have focused on broadly neutralizing monoclonal antibodies elicited by natural infection in humans^22^, polyclonal sera derived from vaccinated humanized cattle^23,24^, and DNA-based vaccination platforms^25^. We therefore sought to investigate the kinetics and durability of the antibody response to PUUV infection using longitudinal sera sampling.

## Methods

### Biospecimen sampling

A total of 23 patients suffering from PUUV infection (confirmed by PUUV IgM point-of-care testing, Reagena POC PUUMALA IgM, and PUUV reverse transcription PCR) were enrolled in the study and described previously ^16,26^. Of these patients, 22% (n=5) exhibited a severe course of disease (defined by the requirement of oxygen, hemodialysis, or intensive care unit admission) and one patient died. The demographic and clinical characteristics of this cohort are described in detail elsewhere^16^. Serum was sampled from these patients at four time points post-hospitalization which cover the early acute (T1, febrile phase, mean sampling time: six days post-symptom onset (PSO), range: 2-11 days PSO), late acute (T2, hypotensive/oliguric phases, mean sampling time: 12 days PSO, range: 8-20 days PSO), early convalescent (T3, polyuric/convalescent phases, mean sampling time: 28 PSO, range: 21-64 days PSO), and late-convalescent (T4, mean sampling time: 197 days PSO, range: 180-256 days PSO) stages of infection (**Fig. 1**). Of the 23 patients enrolled in the study, 20 had serum sampled at T1-4, one patient had samples taken at T1 and T2, and two patients had sera taken only at T1. This study was approved by the institutional review board of the Medical University of Graz (approval no. 33-329 ex 20/21). Written informed consent was obtained from all participants.

**Figure 1.**
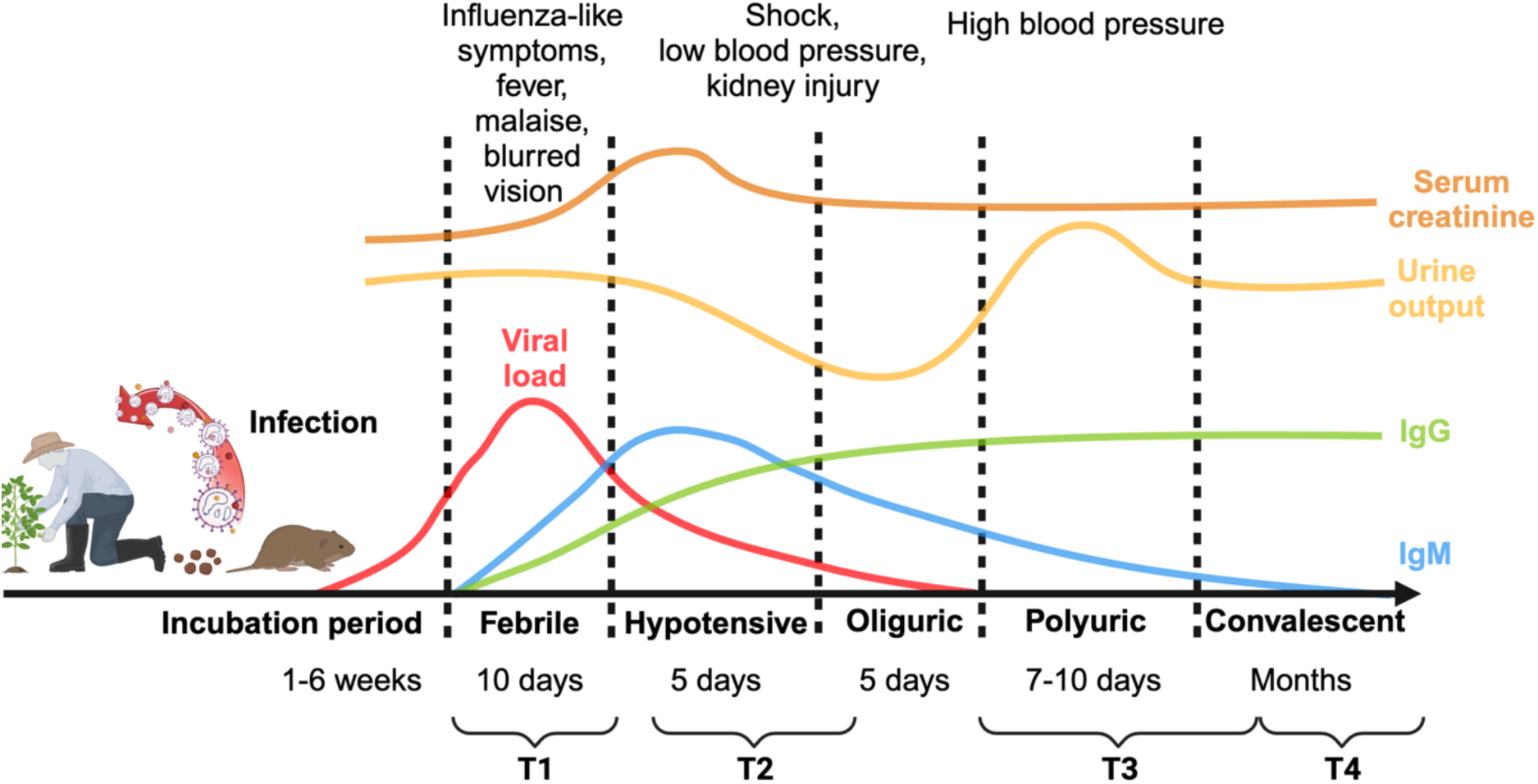
– Schematic timeline of the symptoms associated with HFRS caused by PUUV infection. Following an incubation period of 1-6 weeks, HFRS symptoms manifest as five distinct phases: febrile, hypotensive, oliguric, polyuric, and convalescent. The average time for which each stage lasts is shown, in addition to the symptoms associated with each phase. Serum creatinine, a measure of kidney injury, is displayed, as is urine output as a marker of kidney function. IgM and IgG titers are also shown. The four time points at which patient sera were sampled are also shown in relation to each disease phase. Time point 1 (T1) was sampled six days post-symptom onset (PSO), range: 2-11 days PSO). Time point 2 (T2) was sampled 12 days PSO (range: 8-20 days PSO). Time point 3 (T3) was sampled 28 PSO (range: 21-64 days PSO). Time point 4 (T4) was sampled 197 days PSO (range: 180-256 days PSO). Adapted from Avsic-Zupanc et al.^69^. Created in BioRender. Clark, J. (2025) https://BioRender.com/p36u208

### Cells and viruses

Vero.E6 (American Type Culture Collection, ATCC, CRL-1586, clone E6) and Human hepatocarcinoma Huh.7.5.1 cells (Huh7.5) were cultured in Dulbecco’s modified Eagle medium (DMEM; Gibco), supplemented with Antibiotic-Antimycotic (100 U/ml penicillin–100 μg/ml streptomycin–0.25 μg/ml amphotericin B; Gibco), and 10% fetal bovine serum (FBS; Corning) in a 37°C incubator with 5% CO_2_. rVSVs expressing enhanced green fluorescent protein (eGFP) and the GnGc from HTNV, SEOV,DOBV, and SNV have been previously described^22^, as has the rVSV expressing the PUUV GnGc in addition to a phosphoprotein fused to mNeonGreen (mNG)^27^. Wild-type (WT) VSV (Indiana stain) was obtained from ATCC (VR-1238). The rVSV expressing the GnGc of ANDV is also described elsewhere^28^. Stocks of rVSV-ANDV, -HTNV, -SEOV, -DOBV, and WT-VSV were generated by infecting confluent monolayers of Vero.E6 cells at a multiplicity of infection (MOI) of 0.01 using 1× minimal essential medium (MEM; Gibco) supplemented with 2% FBS, while stocks of rVSV-PUUV and -SNV were generated utilizing Huh7.5 cells and identical methodology. All viruses were grown for 3 days at 37°C before clarification via centrifugation at 4,000g for 10 minutes. At 4°C. Viral stocks were titered using both the 50% tissue culture infectious dose (TCID_50_) method and via classic plaque assay and were stored at −80°C before use. For all infectious assays, Vero.E6 cells were used in conjunction with rVSV-ANDV, -HTNV, -SEOV, -DOBV, and WT-VSV while Huh7.5 cells were used with rVSV-PUUV and -SNV.

### Enzyme-linked immunosorbent assay (ELISA)

To generate antigen for use in ELISA assays, rVSVs were used to infect confluent monolayers of either Vero.E6 or Huh7.5 cells in nine T175 tissue culture flasks (Corning) at an MOI of 0.01. Media was drained from the flasks and replaced with virus diluted in 10 ml of MEM (Gibco) supplemented with 2% FBS. One hour post-infection, the inoculum was removed and replaced with 20 ml MEM (Gibco) supplemented with 2% FBS. Three days post-infection, supernatants were harvested and clarified via centrifugation at 4,000xg for 10 minutes. A 5 ml 30% (w/v) sucrose cushion formulated in 1× NTE buffer (1M NaCl, 100 mM Tris-HCl, and 10 mM EDTA, pH 7.4) was then added to 30 ml of supernatant. Supernatants were then concentrated via high-speed centrifugation at 25,000 rpm at 4°C for 2 hours. The resulting virus pellet was resuspended in PBS (Gibco, pH 7.4) and the protein quantity was determined via Bradford assay (Thermofisher). To generate recombinant Gn proteins, the coding sequences of the PUUV strain Sotkamo (NCBI Reference Sequence: NP_941983.1) and SEOV strain Baltimore (GenBank: AMQ36103.1) Gn ectodomains were cloned into the pFastBacDual (Gibco) vector as previously described^29^. Recombinant baculoviruses were rescued using a published protocol^30^, and purified as described previously^29^. Ultracentrifuge-purified rVSV was diluted in PBS to a concentration of 5µg/ml and coated on Immulon 4 HBX 96-well plates (Thermo Scientific) using 50 μl per well while recombinant Gn proteins were coated at a concentration of 2μg/ml. 24 hours later, the coating solution was removed, and plates were blocked with PBS containing 0.01% Tween-20 (PBST; Fisher Scientific) and 3% non-fat milk (Life Technologies). The blocking solution was incubated for 1 hour before 3 washes using PBST. Serum samples were serially diluted 1:3 from a starting dilution of 1:40 in PBST supplemented with 1% non-fat milk before addition to the respective plates. Sera dilutions were not added to 16 wells per ELISA plate to serve as a blank reading. After a one-hour incubation at room temperature, the plates were washed 3 times with PBST and, when assaying total IgG, 100 μl of mouse anti-human IgG conjugated to horseradish peroxidase (HRP; Sigma-Aldrich, A0293-1ML) diluted 1:3000 in PBST supplemented with 1% non-fat milk was added to each well. When assaying other immunoglobulin subtypes, anti-human IgA HRP (Sigma-Aldrich, A0295, 1:3000), anti-human IgM HRP (SouthernBiotech, 2020-05, 1:3000), anti-human IgG1 (Southern Biotech, 9054-05, 1:5000), anti-human IgG2 (Southern Biotech, 9060-05,1:5000), anti-human IgG3 (Southern Biotech, 9210-0, 1:5000), or anti-human IgG4 (Southern Biotech, 9200-05, 1:5000) were used. Plates were incubated for 1 hour at room temperature, washed three times with PBST, and 100μl of o-phenylenediamine dihydrochloride (Sigma-Aldrich; OPD) was added. Plates were developed for 10 minutes after which the reaction was stopped by adding 50μl of 3M hydrochloric acid (HCl). The plates were read using a Synergy 4 (BioTek) plate reader at an optical density of 490 nanometers. The area under the curve (AUC) for each plate was calculated using GraphPad Prism 10, with the average plus 3 standard deviations of the blank wells serving as the baseline. 12 age-and sex-matched healthy serum controls obtained from the Mount Sinai Health System were utilized as negative controls. The limit of detection for the assay was formulated by taking the average AUC plus three standard deviations of the negative serum samples. An in-house, cross-binding, anti-PUUV human antibody, Ab523, was utilized as a positive control.

### Microneutralization assays

Vero.E6 or Huh7.5 cells were seeded in a 96-well cell culture plate (Corning; 3340) at a density of 2×10^4^ cells per well. Sera was serially diluted 1:3 from a starting dilution of 1:10 to a final dilution of 1:7,290 in 1× minimal essential medium (MEM; Gibco) supplemented with 2% FBS. As with the ELISA assays, the same 12 age- and sex-matched healthy serum controls, and the in-house antibody Ab523, were utilized as negative and positive controls, respectively. 24 hours later the rVSV was diluted to 10^4^ tissue culture infectious dose 50 (TCID_50_)/mL, and 80 μL of virus and 80 μL of sera dilution were combined and incubated together for 1 hour at room temperature on a separate 96-well plate. After the incubation, 120 μL of virus–antibody inoculum was used to infect cells for 1 hour at 37°C. The inoculum was then removed and 100 μL of each corresponding sera dilution was added to the wells in addition to 100 μL of 1X MEM supplemented with 2% FBS. Throughout this process, a total of 6 wells per plate were incubated with 1xMEM 2% FBS containing no virus or sera, and another 6 wells with 1xMEM 2% FBS containing virus with no sera to serve as control wells for the quantification of % inhibition. Cells were incubated at 37°C in a 5% CO_2_ incubator for 2 days and fixed with 10% paraformaldehyde (Polysciences) for 24 hours at 4°C. The paraformaldehyde was removed, cells were washed twice with PBS, and permeabilized via the addition of 100µl of PBS supplemented with 0.1% Triton X-100 (Fisher). After 15 minutes, the Triton X-100 solution was removed, and cells were blocked with PBS supplemented with 3% non-fat milk (American Bio; AB10109-01000) for one hour at room temperature. The cells were stained using a mouse anti-VSV-N antibody (Kerafast, clone 10G4) diluted 1:3000 in PBST 1% milk for one hour. Cells were then washed twice with PBS and anti-mouse IgG conjugated to horseradish peroxidase (Rockland, 610-603-002) diluted 1:3000 in PBST 1% milk was added to each well. After one hour, the plates were washed twice using PBS and developed via the addition of 100μl of o-phenylenediamine dihydrochloride (Sigma-Aldrich; OPD). Ten minutes later, 50μl of 3M hydrochloric acid (HCl) was added to stop the reaction, and the plates were read using a Synergy 4 (BioTek) plate reader at an optical density of 490 nanometers. The 50% inhibitory dilution (ID_50_) of each serum sample was calculated using a previously described methodology^31^.

### ADCC *in vitro* reporter assays

Huh7.5 cells were seeded in white 96-well microplates (Sigma-Aldrich) at a density of 2×10^4^ cells per well. The next day, cells were infected with rVSV-PUUV at an MOI of 1 diluted in 1x MEM supplemented with 2% FBS, 100 μl per well, for one hour. After one hour, the inoculum was removed and replaced with 100 μl 1x MEM 2% FBS. 48 hours later, patient sera were serially diluted 1:3 from a starting dilution of 1:10 in 1× MEM supplemented with 2% FBS to a final dilution of 1:21,870. The same age- and sex-matched healthy serum controls utilized in the ELISAs and microneutralization assays were again employed as negative controls, and the in-house antibody, Ab523, was again utilized as a positive control. The media on the wells was aspirated and replaced with 25 μl Roswell Park Memorial Institute (RPMI-1640) media (Thermofisher) containing 2% low-IgG FBS (Promega) and 25 μl of sera dilution. A total of 16 wells did not have sera added to them to serve as background luminescence readings. As with the ELISAs and microneutralization assays, 12 age- and sex-matched healthy serum controls, and the in-house antibody Ab523, were utilized as negative and positive controls, respectively. 3×10^6^ ADCC Bioassay Effector Cells expressing the human FcγRIIIa receptor (Promega) diluted in 25 μl RPMI-1640 2% low-IgG FBS were added per well and the cells were incubated at 37°C with 5% CO_2_ for six hours. 75 μL of Bio-Glo luciferase reagent (Promega) was added to each well and luminescence was quantified using a Synergy 4 (BioTek) plate reader. The 16 wells with no added sera were used to determine a background luminescence reading which was used to calculate the AUC using GraphPad Prism 10.

### Cytokine analysis

The concentrations of IL-18, IL-33, MCP-1, IL-8, IFNα, IFNβ, IFNγ, IL-13, IL-23, IL-1β, IL-22,IL-6, IL-4, IL-5, GM-CSF, IL-12p70, IL-10,TNFα, IL-17F, IL-2 and IL-17A in serum samples were determined using LEGENDplex bead-based immunoassay (Biolegend, USA) in accordance with the manufacturer’s instructions. Briefly, 25 μL of serum was mixed with equal volumes of assay buffer, and beads to bring the final volume to 75 μL per well. Serial dilution of standards was prepared in the assay buffer and further processed in the same way. The samples were incubated in the dark on a plate shaker at 750 rpm for two hours at room temperature. After washing with 200 μL of wash buffer (centrifuged at 1100 rpm for 5 minutes at 25°C), the samples were incubated with 25 μL of detection antibodies for one hour on a plate shaker. Following this, an equal volume of Streptavidin - R-phycoerythrin conjugates was added, and the samples were incubated for another 30 minutes with subsequent washing and reconstitution in 100 μL of PBS. Data were collected using the Attune flow cytometer (Thermo Fisher Scientific) and processed with FlowJo_v10.8.1 and LEGENDplex™ data analysis software (https://legendplex.qognit.com).

### Statistical analysis

For ELISA, microneutralization, and ADCC reporter assay data, statistical analysis was performed in GraphPad Prism. AUC and ID_50_ values were log10 transformed and statistical significance was interrogated via ordinary one-way ANOVA using a Tukey’s multiple comparisons test. Significance was defined as p<0.05 and is indicated in graphs where present. Cytokine concentrations in serum samples from PUUV-infected patients at various time points were compared using the pairwise Wilcoxon test. Normality of sample distribution was assessed with the Shapiro-Wilk test. Pearson or Spearman correlation analyses were applied to normally or non-normally distributed variables, respectively, with p-values adjusted for multiple comparisons using the Benjamini-Hochberg FDR method. For heatmap visualization, a log(p+1) transformation was applied, followed by min-max normalization. Ward’s clustering algorithm was employed to group individuals based on similar immune response characteristics. Comparisons between groups identified through serum cytokine profiles were conducted using Student’s t-test. Statistical significance was defined as p < 0.05. All analyses were performed in RStudio (R version 4.3.3) and Google Colab (Python version 3.10).

## Results

### Infection elicits a strong anti-PUUV IgG response with cross-binding antibodies produced at convalescence

To investigate the humoral response to PUUV infection, we utilized a previously described cohort of patients from Graz, Austria, who were hospitalized due to PUUV infection and had longitudinal sera collected not only during hospitalization but also after discharge (up to 256 days after symptom onset)^16^. Sera sampling covered the five phases of disease progression (**Fig. 1**). To assess the IgG response to PUUV, we utilized ultra-centrifuge purified recombinant vesicular stomatitis virus (rVSV) expressing the full-length GnGc of PUUV as antigen for use in enzyme-linked immunosorbent assays (ELISAs)^22^. Each of the serum samples was assayed for the presence of anti-PUUV IgG alongside 11 age- and sex-matched healthy serum controls obtained from the Mount Sinai Health System. Due to serum scarcity, some samples were prioritized for microneutralization assays instead of ELISAs. The samples that were assayed using each technique is shown in **Table. S1**. At T1, 17 of the 21 serum samples (80.9%) exhibited a robust anti-PUUV IgG response which was maintained at T2 and T3 at which points 19 of the 21 serum samples (90.5%) displayed IgG (**Fig. 2A**). The IgG titer remained high at T4, 6-7 months after infection at which point all the patients (20/20) had seroconverted, indicating a strong adaptive immune response to infection. To determine whether cross-binding antibodies are elicited by PUUV infection, IgG ELISAs were performed using rVSVs expressing the GnGc of several phylogenetically and antigenically distinct hantaviruses, including SEOV, HTNV, DOBV, ANDV, and SNV. Anti-SEOV IgG was absent from most patients at T1-3 however, titers were detected at T4 for 50% of patients (**Fig. 2B**). Similarly, anti-DOBV titers were low at T1-3 but detectable in 30% of serum samples at T4 (**Fig. 2C**). Conversely, more patients exhibited detectable anti-HTNV IgG titers at T1-3, with 13 of 21 patients (65%) displaying IgG at T4 (**Fig. 2D**). Anti-SNV IgG titers were low at T1-3 and detectable only in 10% of serum samples at T4 (**Fig. 2E**). A small number of patients also exhibited anti-ANDV IgG at T 1-3, however by T4, 15 of the 20 patients (75%) had detectable IgG titers (**Fig. 2F**). This indicates that cross-binding antibodies are elicited by PUUV infection, with the highest proportion of patients at T4 exhibiting cross-reactive IgG, but at early time points this varied according to the hantavirus GnGc tested. To verify that the observed IgG responses are hantavirus-specific and not due to antibodies binding to the rVSV vector, ELISAs were carried out using wild-type VSV. IgG titers were comparable between the PUUV patient sera and control sera, indicating that the observed IgG titers are specific to the hantavirus glycoproteins tested (**Fig. 2G**). Most serum samples exhibited a high background AUC as determined by the average plus three times the standard deviation of the negative control samples. To determine if the high background values were due to the use of rVSV pseudo particles, we employed recombinant PUUV and SEOV Gn for comparison with the full-length GnGc expressed by the rVSVs. Titers against PUUV Gn were reduced compared to the full-length GnGc (**Fig. S1A**), while titers against SEOV Gn were comparable (**Fig. S1B**). The limit of detection was slightly smaller for the recombinant Gn ELISA compared to the GnGc, therefore, more patients exhibit detectable IgG titers at T4 (12/20, 60%) compared to rVSV-SEOV ELISA (10/20, 50%).

**Figure 2.**
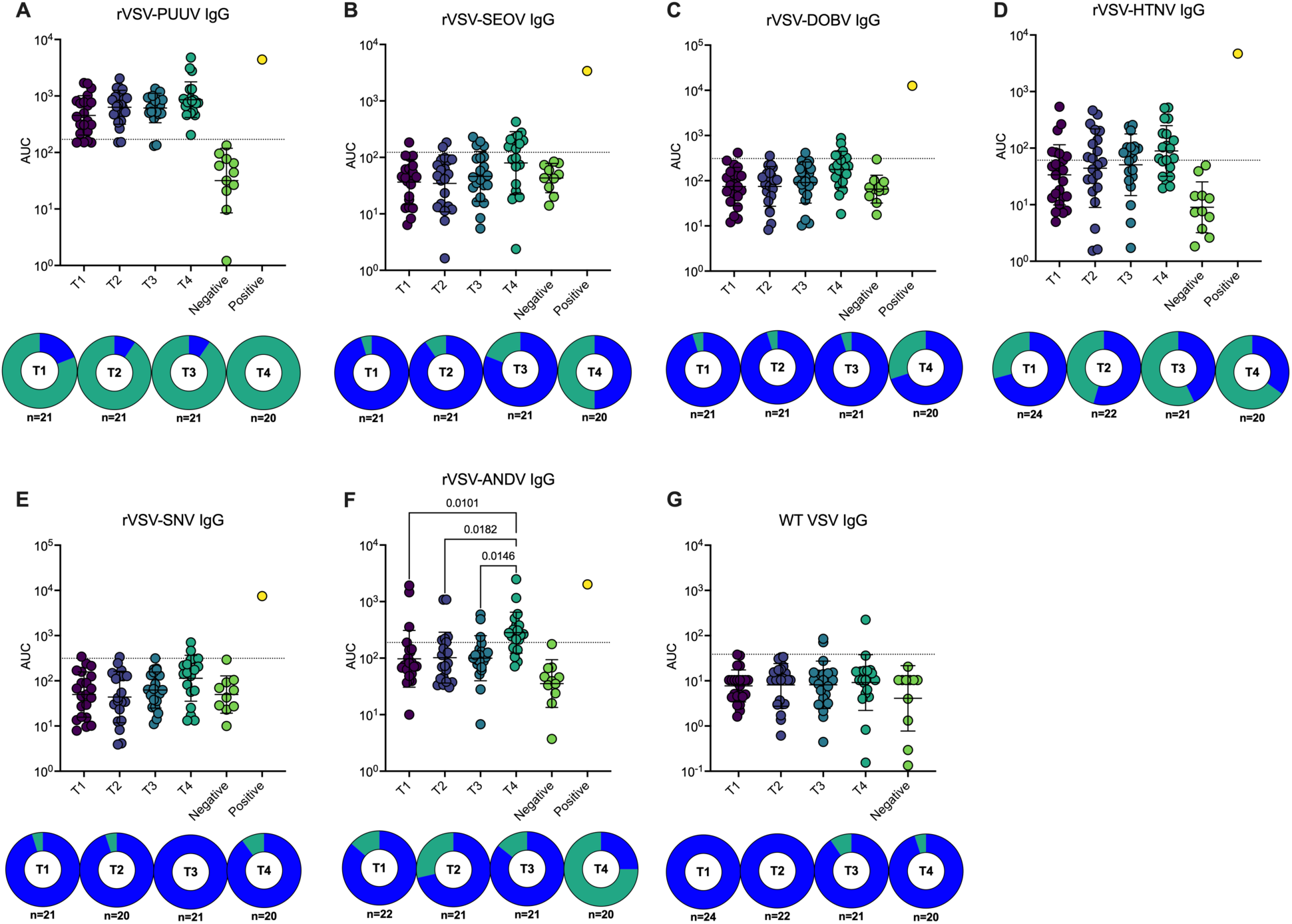
– Patient sera exhibit robust anti-PUUV IgG at all time points and cross-binding IgG by time point 4. ELISAs were carried out using patient sera and plates coated with ultracentrifuge purified rVSV expressing the GnGc of **A)** PUUV, **B)** SEOV, **C)** DOBV, **D)** HTNV, **E)** SNV, or **F)** ANDV. Wild-type VSV **G)** was used as a negative control. IgG titers are shown as area under the curve (AUC) with the geometric mean and geometric standard deviation denoted by error bars. 11 age and sex matched healthy control sera were included as negative controls and an in-house, anti-PUUV monoclonal antibody was used as a positive control. AUC values were log10 transformed and statistical significance was interrogated via ordinary one-way ANOVA using a Tukey’s multiple comparisons test. Only comparisons with p ≤ 0.05 are shown. Limits of detection (LOD) were calculated for each antigen using the average + 3x standard deviations of the AUC of the negative control sera and are shown as dotted lines on each graph. The percentage of serum samples with LODs above the limit of detection are shown as pie charts, with the percentage of positive samples displayed in green. The n for each time point is displayed below each pie chart.

### Neutralizing activity is detected at early time points and is highest at convalescence

As PUUV infection elicits a strong IgG response, with samples taken at T4 exhibiting cross-binding antibodies, we utilized the rVSV pseudoviruses for microneutralization assays to determine whether these sera demonstrated neutralizing activity. Like the IgG ELISA data, patients exhibited robust anti-PUUV neutralizing titers at early time points which were significantly higher at T4 compared to T1 (**Fig. 3A**). Conversely, despite most patients displaying little detectable IgG at T1-3, some patient sera neutralized rVSV-SEOV at earlier time points, with 95% (19/20) of patients showing neutralizing activity at T4 (**Fig. 3B**). Similarly, although most patients did not exhibit anti-DOBV IgG titers at both acute and convalescent time points, neutralizing titers were detected at T1-3 with 95% (19/20) of T4 sera displaying detectable neutralization (**Fig. 3C**). A higher proportion of patients displayed neutralizing titers against rVSV-HTNV in 75% (18/24) of T1 sera, 86.4% (19/22) at T2, 66.7% (14/21) at T3, and finally 85% (17/20) at T4 (**Fig. 3D**). Neutralizing activity against rVSV-SNV was more muted, with a small minority of patient sera at T1-3 exhibiting neutralizing titers which then increased to 30% (6/20) at T4, though to a greater extent than the IgG titers (**Fig. 3E**). Again, this is unlike the ELISA data in which a smaller proportion of patients had detectable anti-HTNV IgG which peaked at T4. Finally, rVSV-ANDV neutralizing titers were detected in 42.7% (10/24) patients at T1 and 54.2-52.4% (13/24 – 11/21) at T2-3 before increasing to 75% (15/20) patients at T4 (**Fig. 3F**). Again, wild-type VSV was utilized as a negative control to ensure that the observed neutralization is due to the specific neutralization of the hantavirus GnGcs expressed by each pseudovirus. Like the ELISA data, none of the patient sera neutralized wild-type VSV, indicating that the observed neutralization is due to anti-GnGc antibodies (**Fig. 3G**).

**Figure 3.**
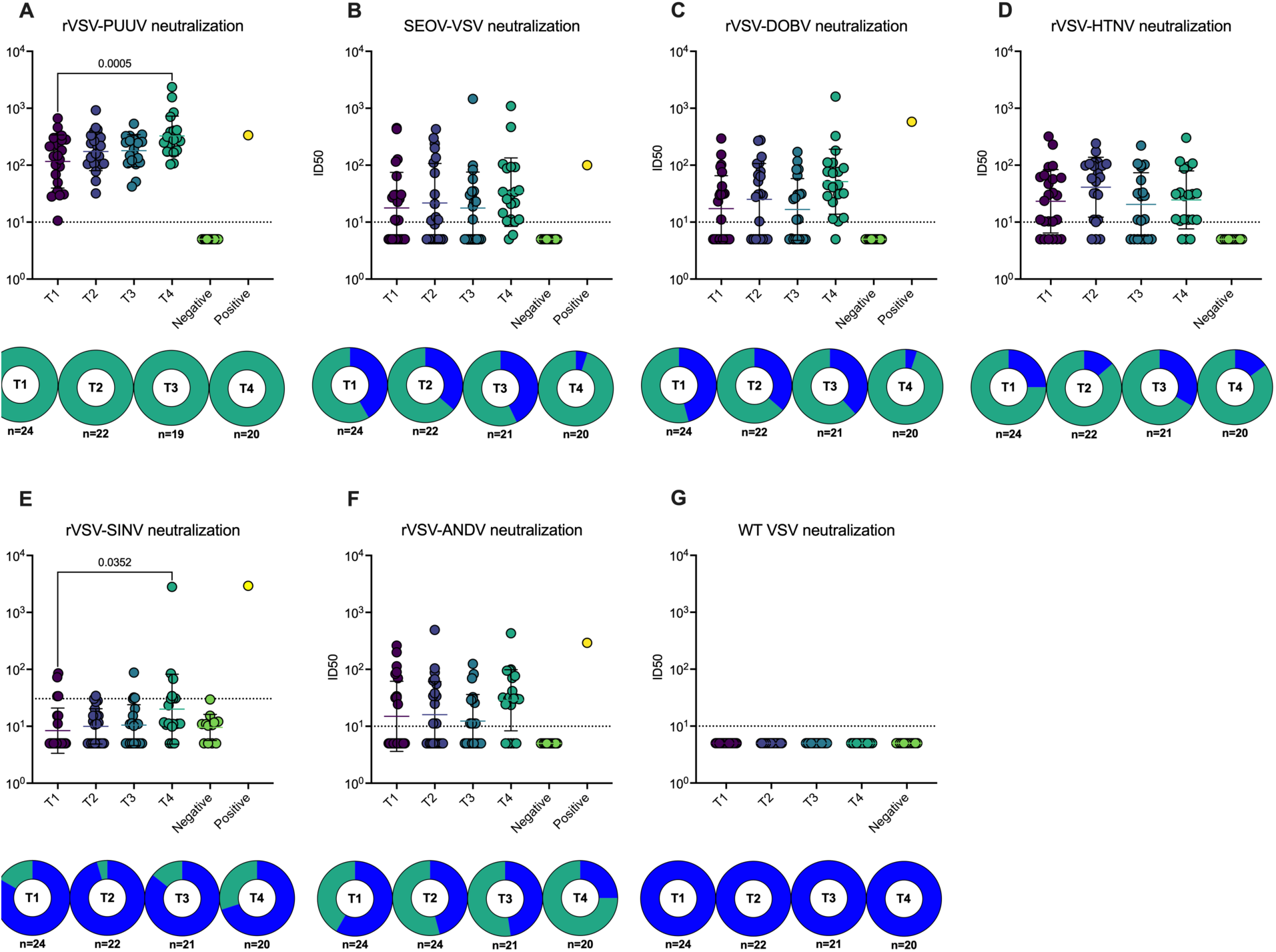
– Some patient sera exhibit cross-neutralization at acute and early convalescent time points, with the highest proportion of sera with neutralizing activity at T4. Microneutralization assays were carried out using patient sera and rVSV pseudoviruses expressing the GnGc of **A)** PUUV, **B)** SEOV, **C)** DOBV, **D)** HTNV, **E)** SNV, or **F)** ANDV. WT-VSV **G)** was utilized as a negative control. Neutralizing titers are shown as 50% inhibitory dilutions (ID_50_s) with the geometric mean and geometric standard deviation denoted by error bars. 11 age and sex matched healthy control sera were included as negative controls and an in-house, anti-PUUV monoclonal antibody was used as a positive control. ID_50_ values were log10 transformed and statistical significance was interrogated via ordinary one-way ANOVA using a Tukey’s multiple comparisons test. Only comparisons with p ≤ 0.05 are shown. Limits of detection (LOD) are shown as dotted lines on each graph. The percentage of serum samples with LODs above the limit of detection are shown as pie charts with the percentage of positive samples displayed in green. The n for each time point is displayed below each pie chart.

To determine whether other immunoglobulin subclasses with cross-binding potential may be present in the patient sera, we performed ELISAs utilizing anti-IgM and IgA secondary antibodies. We found that PUUV GnGc-specific IgM was present in around 39.1% (9/23) of the patients at T1, 40.9% (9/22) at T2 and 33.3% (7/21) at T3. This significantly decreased by T4 with all patients except one exhibiting titers below the limit of detection at T4 (**Fig. 4A**). Anti-PUUV IgA was also detectable in 20.1% (6/23) of patient sera at T1, which increased to 40.9% (9/22) at T2 before moderately decreasing to 35% (7/20) by T4 (**Fig. 4B**). To determine if cross-binding IgM or IgA was elicited in response to infection, we carried out ELISAs using ultracentrifuge-purified rVSV-SEOV. Cross-binding anti-SEOV GnGc IgM was present in 17.4% of patient sera at T1, which increased to 27.3% (6/22) and 25% (5/20) patients by T2 and T3 before decreasing below the limit of detection for all but two patients by T4 (**Fig. 4C**). Anti-SEOV GnGc IgA was present in 29.2% (7/24) of patient sera at T1, and persisted until T2, before reducing below the limit of detection for all but two patients at T3 and T4 (**Fig. 4D**). This indicates that cross-binding IgM and IgA are present at earlier time points and that these may be responsible for the observed cross-neutralization of other hantaviruses before cross-binding IgG titers increase at t4.

**Figure 4.**
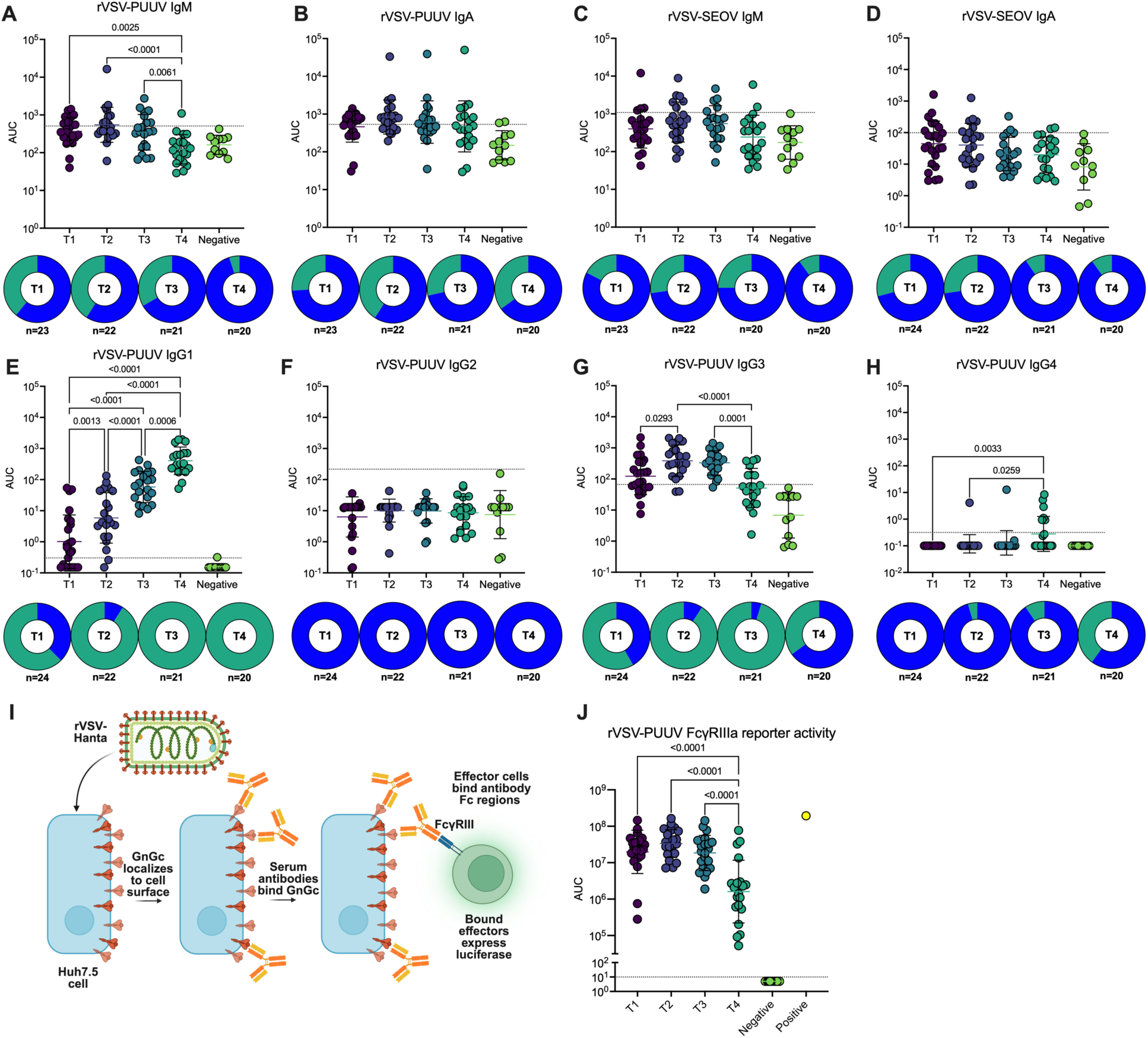
– The quantity of immunoglobulin subtypes changes between the acute and convalescent phases and is associated with a decrease in Fc effector function. ELISAs were carried out using patient sera to investigate **A)** anti-PUUV IgM, **B)** anti-PUUV IgA, **C)** anti-SEOV IgM, and **D)** anti-SEOV IgA. IgG subtype ELISAs were performed to investigate the quantities of anti-PUUV **E)** IgG1, **F)** IgG2, **G)** IgG3, or **H)** IgG4. **I)** A schematic of the luciferase-based reporter assay, which was utilized to characterize the effector activity promoted by patient sera. RVSV-PUUV infected Huh.75 cells were incubated with patient sera and reporter cells that express the FcγRIIIa receptor, coupled to a gene expression pathway that results in the production of luciferase. **J**) The FcγRIIIa activity of the sera re shown as AUCs, with the mean and standard deviation denoted by error bars. 11 age and sex matched healthy control sera were included as negative controls, and an in-house, anti-PUUV monoclonal antibody was used as a positive control. AUC values were log10 transformed and statistical significance was interrogated via ordinary one-way ANOVA using a Tukey’s multiple comparisons test. Only comparisons with p ≤ 0.05 are shown. Limits of detection (LOD) are shown as dotted lines on each graph. The percentage of serum samples with LODs above the limit of detection are shown as pie charts with the percentage of positive samples displayed in green. The n for each time point is displayed below each pie chart.

### The proportion of the IgG subclasses changes post-infection and is associated with a decreased FcγRIIIa activation

As cross-reactive IgG is most abundant in collected sera at T4, we sought to determine whether this increase is associated with changes in the abundance of IgG subclasses. We utilized ultracentrifuge-purified rVSV-PUUV alongside a panel of IgG subclass-specific secondary antibodies to quantify IgG1, IgG2, IgG3, and IgG4 in the patient sera. At T1, anti-PUUV IgG1 was present in 66.7% (16/24) of the patient sera, albeit at low titers (**Fig. 4E**). The titer significantly increased by T2. At that point, 90.9% (20/22) of patients exhibited detectable anti-PUUV IgG1 (**Fig. 4E**). These titers again significantly increased by T3 at which point all the patients exhibited a robust IgG1 response (**Fig. 4E**) which again increased significantly at T4 (**Fig. 4E**). Conversely, IgG2 was absent from all patient sera at all time points (**Fig. 4F**). IgG3 was present in 58.3% (14/24) of the patient sera at T1. It significantly increased at T2 at which point 90.9% (20/22) of patients had detectable titers (**Fig. 4G**). These titers remained constant at T3 however, at T4 the IgG3 titers significantly decreased and were below the limit of detection for 65% of the patients (**Fig. 4G**). IgG4 was absent from all the patient sera at T1 and was detectable in a single patient at T2 and in two patients at T3 (**Fig. 4H**). At T4 there was a significant increase in IgG4, with 40% (8/20) of patients displaying low, but detectable titers (**Fig. 4H**). As the Fc portions of the IgG subtypes interact with the Fcγ receptors expressed on immune cell populations to different extents, we sought to determine if the sera from each time point could activate FcγIIIa. We therefore carried out antibody-dependent cellular cytotoxicity (ADCC) reporter assays using cells infected with rVSV-PUUV (**Fig. 4I**). At T1, all the serum samples exhibited robust FcγIIIa activity, which was maintained at T2-3 (**Fig. 4J**). At T4 however, this activity was significantly reduced by 10-100-fold for most patients, with only 4 patient samples maintaining high levels of activity (**Fig. 4J**). This indicates that changes in the abundance of the IgG subtypes likely influences the Fc-dependent activity of the sera.

### Patients can be segregated based on the serum abundance of Th1 and Th2 associated cytokines

While conducting serology to assess antibody responses, we utilized the same samples to evaluate cytokine responses in the serum of the initial 24 individuals. Concentrations of inflammation type-I/Th1-associated cytokines (IL-1β, IL-6, IL-2, MCP-1, TNFα, IL-8, IL-18, IFNα, IFNβ, IFNγ, IL-12p70), inflammation type-II/Th2-associated cytokines (IL-10, IL-33, IL-13, IL-4, IL-5), and inflammation type-III/Th17-associated cytokines (IL-23, IL-22, GM-CSF, IL-17F, and IL-17A) in serum samples were determined. The concentration of all analyzed cytokines increased in the acute stage (T2) compared to the late convalescent stage (T4). The most significant increases relative to the “baseline” (cytokine concentrations in the convalescent stage) were observed for IFNγ, IL-6, IL-13, IL-10, and IL-17 (**Fig. 5A**). Patients with severe PUUV infection (as defined by a composite endpoint: ICU admission, need for oxygen supply, and/or hemodialysis) exhibited elevated serum concentrations of IL-23, IL-22, IL-4, IL-13, IL-5, IL-2, and IL-10 (**Fig. 5B**). Based on principle component analysis (PCA) results, two major patterns of cytokine expression can be identified: one) IL-2, IL-4, IL-5, IL-10, IL-13, IL-22; two) IL-6, IFNγ, IL-8, IFNα, IFNβ, TNFα, IL-12p70, IL-33, GM-CSF (**Fig. 5C**). The first set of cytokines is primarily represented by major regulators of the type-II inflammatory response, produced by type 2 innate lymphoid cells (ILC-2) and T helper 2 (Th2) cells, while the second set consists of inflammation type-I-associated cytokines. Interestingly, the type-II-polarized immune response was primarily associated with severe disease cases in the acute stage of the infection. The highest correlation coefficients were shown between the concentrations of IFNα, IFNγ, IL-12p70, IL-23, and IL-33 and HTNV-neutralizing antibody titers in the acute phase of the infection (**Fig. 5D**). Additionally, positive correlations were shown between the levels of neutralizing antibodies against ANDV, DOBV, HTNV, and SEOV in the convalescent stage and the concentrations of IL-23 in the acute stage of the PUUV infection (**Fig. 5E**). However, the revealed associations between the analyzed variables were not statistically significant according to the results of Spearman correlation analysis (**Fig. S2**).

**Figure 5.**
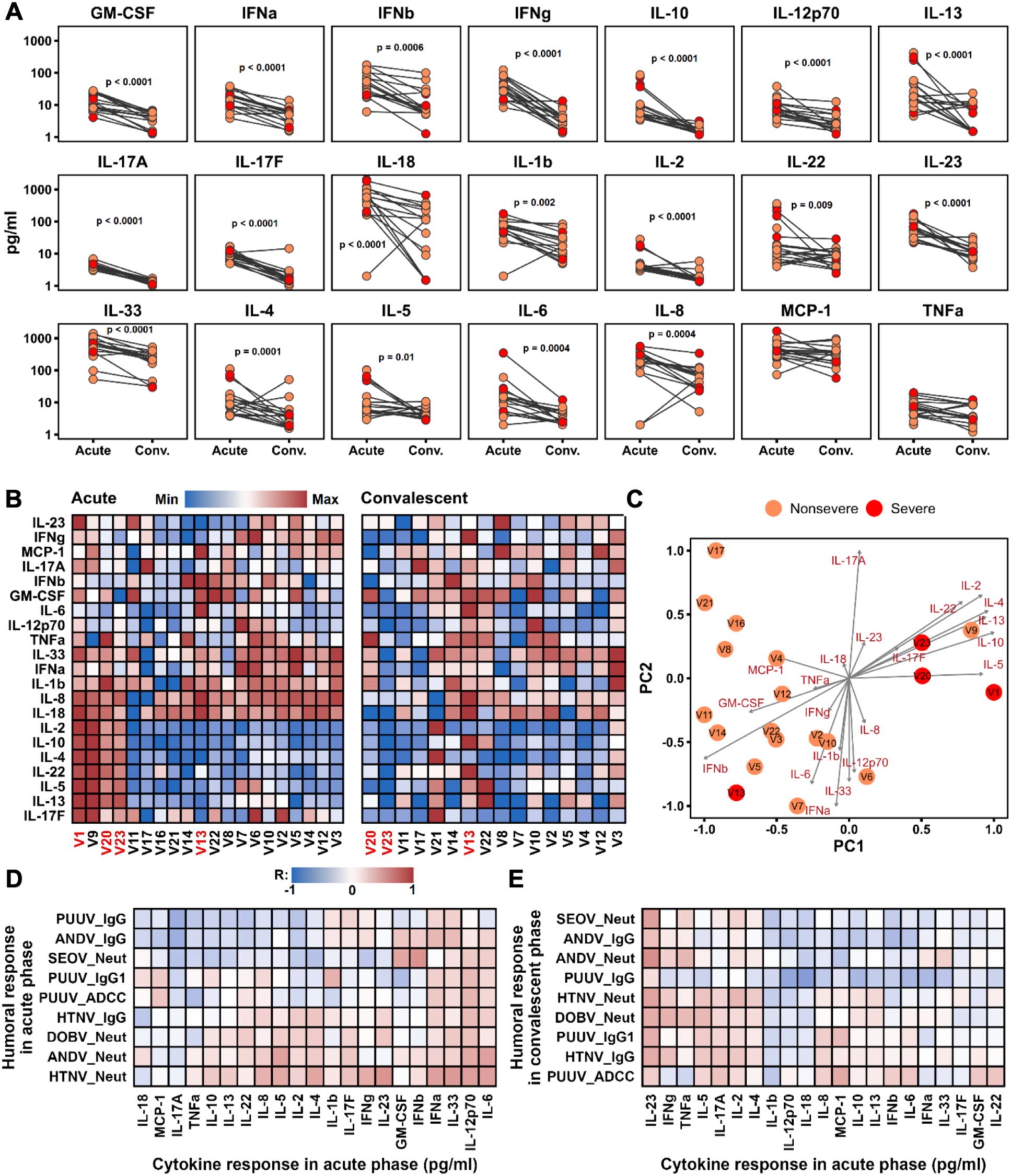
– Cytokine profile or patients at acute and convalescent disease stages. **A)** Concentrations of cytokines in serum samples of PUUV-infected patients at acute (T2) vs convalescent (T4) time points. Values were compared using pairwise Wilcoxon test (*: p < 0.05). **B)** Heat maps display the normalized serum concentration of each cytokine in each sample at the acute (T2) and convalescent (T4) phases. Values were normalized using log(p+1) transformation and min-max normalization. Severe disease cases are indicated with red labels. **C)** Principal component analysis (PCA) biplots representing the distribution of PUUV-infected patients by the serum concentration of cytokines at T2 as well as factor loads of analyzed variables on the first two principal components. Severe cases are shown in red. **D-E)** Heatmaps represent Spearman correlation analysis between acute-phase cytokine concentrations and humoral immune response parameters in the acute phase **(D)** as well as between acute-phase cytokine concentrations and humoral immune response parameters in the convalescent phase **(E)**. Only humoral immune response variables with a seroconversion rate exceeding 50% were included in the correlation analysis. To refine the data, the limit of detection (LOD), calculated as three times the standard deviation (SD) of negative samples, was subtracted from the values used in the analysis. Any resulting negative values were replaced with zero. Log(p+1) transformation and min-max normalization were applied to the original data. Benjamini-Hochberg FDR adjustment was used for multiple comparisons.

## Discussion

Investigating the humoral immune response, we found that PUUV infection resulted in a strong IgG response, which was detectable in the patients at T1, shortly after disease onset. This has been previously reported and is characteristic of the long incubation period associated with PUUV infection, which ranges from 2-6 weeks^13,32,33^. Anti-PUUV total IgG titers were stable from T1 to T4, at which point all the patients exhibited detectable IgG. Interestingly, cross-binding antibodies capable of binding the GnGc of both old-world (SEOV, HTNV, DOBV) and new-world hantaviruses (SNV, ANDV) were detectable in a subset of patients, predominantly at T4. The appearance of cross-binding IgG, specifically 6 months post-infection, indicates that affinity maturation favoring conserved epitopes shared amongst phylogenetically distinct hantaviruses takes place in the months following PUUV infection. A potent anti-PUUV neutralizing antibody response was also noted, with sera from all time points exhibiting neutralizing activity, which peaked at T4. Unlike the IgG ELISA data, over 50% of patients displayed cross-neutralizing activity against rVSV-SEOV, -DOBV, and -ANDV at T1-3, which increased to over 75% of patients at T4. Patient sera showed a more muted cross-neutralizing response against rVSV-SNV and an enhanced response against rVSV-HTNV which more closely matched the observed anti-SNV and anti-HTNV IgG titers. For each of the VSV pseudoviruses tested, the observed number of serum samples that displayed cross-neutralizing activity was higher than those that demonstrated cross-binding IgG. These results could also be partially explained by differences in sensitivity between the ELISA and microneutralization assay methodologies, particularly due to the high background IgG values noted in the age and sex-matched healthy donors which were utilized as negative control samples. This discrepancy at earlier time points could be explained by the presence of cross-neutralizing IgM and IgA. Indeed, cross-binding anti-SEOV IgM was detectable in patients at T1 and peaked and remained stable at T2-3 before subsiding by T4 while IgA was detectable in around a third of patients at T1 and T2 before clearing by T3. While immunoglobulin subclasses aside from IgG exert neutralizing activity, we are unable to determine which of these subclasses may be responsible for the observed cross-neutralization utilizing only ELISA. This question could be answered by depleting IgG from the sera before carrying out additional neutralization assays. However, due to sera scarcity, we were unable to utilize this methodology.

It is interesting that cross-reactive IgG titers are highest at T4, 6-7 months after disease onset. These patients are presumably exposed to the virus a single time, however, months after infection IgG class switching takes place giving rise to increased IgG1, decreased IgG3, and increased IgG4 concurrent with increasing titers of cross-binding antibodies. The development of IgG4 antibodies in response to infection is poorly understood, however, it is associated with repeated or chronic exposure to antigens. Repeated SARS-CoV-2 mRNA vaccination has been shown to induce IgG4 responses^34^, as has repeated exposure to phospholipase from honey bee stings^35,36^. Similar changes in IgG repertoire have also been demonstrated in survivors of Ebola virus (EBOV) infection^37^, with strong evidence for persistent viral infection in immune-privileged tissues such as the eyes and testes.^38–42^. Repeated exposure to antigen appears to be necessary but not sufficient to induce IgG class-switching towards IgG4, as repeated tetanus vaccination has not been found to stimulate an IgG4 response^43^, nor was persistent infection with human cytomegalovirus^44^. Interestingly, increased IgG4 titers have been identified in one previous publication in which anti-Gn and anti-Gc IgG4 titers were found to be significantly increased 2 years post PUUV infection^45^. This study also noted an increase in IgG1 titers and a decrease in IgG3 titers, in line with our findings^45^. These data suggest that PUUV may establish persistent or latent infections in humans, or that viral antigen is somehow retained and continuously presented to the immune system. Indeed, neutralizing antibody titers against PUUV have been shown to increase in the years after infection, with some patients exhibiting high titers decades after infection^46^. Additional sera sampling from the cohort described here one-to-two years post infection would shed further light on this phenomenon. Other evidence for hantavirus persistence in humans comes from studies that have shown the presence of high levels of highly differentiated long-lived CD127^-^ memory T cells years after ANDV infection^47^, and the expansion of NK cells that remained functional over 60 days after PUUV infection^48^. In contrast to PUUV infection, IgG1 has been reported as the predominant subclass in convalescent Choclo virus (CHOV) patients^49^, while IgG3 has been identified as dominant in convalescent SNV patients^50^. However, in the absence of ongoing class switching, ANDV and SNV patients also exhibit elevated neutralizing antibody titers years after exposure^51,52^. Additional experiments, including postmortem tissue analysis, must be carried out to determine the possible location of viral persistence before it can be confirmed that hantaviruses establish persistent infections in humans.

Presently, no therapeutics are available for the treatment of hantavirus infections, and current treatment strategies are limited to the alleviation of disease symptoms. The broad-acting antiviral ribavirin is effective in the treatment of HTNV infection if given soon after symptom onset^53,54^, however, there is limited evidence of its efficacy in the treatment of HCPS^55,56^, and it failed to reduce PUUV viral loads in a clinical trial^57^. Conversely, convalescent plasma from survivors of ANDV infection is efficacious in the treatment of HCPS^58^. Therefore, one promising therapeutic avenue is the use of human-derived monoclonal antibodies cloned from the B cells of hantavirus infection survivors. Potently neutralizing antibodies cloned from convalescent ANDV patients are protective in the Syrian golden hamster challenge model^59^, promoting partial protection even when administered 10 days post-infection^60^. Broadly neutralizing antibodies have also been identified in survivors of SNV infection and PUUV infection^22,61^. Our results indicate that the optimal time point for the cloning and production of broad-binding antibodies is upwards of 6 months post-infection. The results presented in this study should, therefore, be taken into consideration in future studies focused on the identification of therapeutic, human-derived, monoclonal antibodies.

PUUV-infected patients were characterized by a substantial increase in the production of proinflammatory and anti-inflammatory cytokines associated with all major branches of the immune response (Th1, Th2, Th17). The most significant increase was shown for IFNγ, IL-6, IL-13, IL-10, and IL-17. Elevated levels of IL-6, IL-10, and IFN-γ have previously been reported in the sera of acute PUUV patients^62^, severe HCPS cases^63^, and PUUV-infected cynomolgus macaques^64^.

Patients with severe PUUV infection were characterized by the elevated production of Th2/Th17-associated cytokines IL-23, IL-22, IL-4, IL-13, and IL-5. This suggests a potentially dysregulated immune response contributing to disease severity. It was shown previously that the protective immune response to hantaviral infections is mediated primarily by Th1 CD4+ and cytotoxic CD8+ T-cells^65,66^. Patients with severe/critical HFRS showed lower amounts of single cytokine (IFN-γ, IL-2, and TNF-α) or dual-cytokine-producing CD4+ Th1 cells. On the other hand, a trend towards higher levels of perforin+ CD4+ T cells was observed in the mild/moderate group. Th2 responses usually counterbalance Th1 inflammation and excessive Th2 activation might not effectively control the virus and could lead to immunopathology ^67^. It has previously been demonstrated that ILC2, which are major regulators of type II inflammation and are responsible for Th2 polarization, play a significant role during the acute phase of PUUV infection^68^. Viral infection induces the release of alarmins such as IL-25, IL-33, and thymic stromal lymphopoietin (TSLP) which activate ILC2 thereby promoting the secretion of IL-4, IL-5, and IL-13. This increased production of type-II-associated cytokines was also shown to be associated with higher disease severity^68^. The elevated levels of IL-23 in the sera of acute patients have been observed in other studies, however, we demonstrated a non-statistically significant correlation between the IL-23 levels in the acute stage of infection and the cross-neutralizing humoral response against various hantaviruses (ANDV, DOBV, HTNV, SEOV) in the convalescent stage. This suggests that, while IL-23 may contribute to acute severity, it also promotes broader antibody-mediated immunity over time. Taken together, these data identify biomarkers of PUUV infection which may have diagnostic utility and may inform the analysis of future hantavirus vaccine design platforms that aim to promote broad antibody responses to protect against multiple hantaviruses.

## Contributions

SH and RK performed clinical work in Austria. JC, KV, RAS, JSY performed assays. EM, EK and KC provided reagents. RK, VS and FK supervised the study. JC, SH, RK and FK developed the concept. JC, KV and SH drafted the manuscript. JC, KV, SH, VS, RK and FK performed the data analysis. All authors edited and approved the manuscript.

## Data availability statement

All data is available from ImmPort xxx.

## Acknowledgments

We would like to thank Dr. Charlie Rice for kindly providing the Huh7.5 cells. We would like to thank Dr. Taia Wang for her advice on IgG subtype secondary antibody purchasing. We thank Sean Whelan (Washington University) for providing the VSV-PUUV used in this study. The study was supported by a research grant of the Austrian Science Fund (FWF) to SH (number J 4737-B). The study was also supported by the Styrian government (project no. ABT12-106729/2022-13 to RK). Work in the Krammer laboratory at the Icahn School of Medicine at Mount Sinai was supported by institutional funding.

---

**Supplementary Figure S1.**
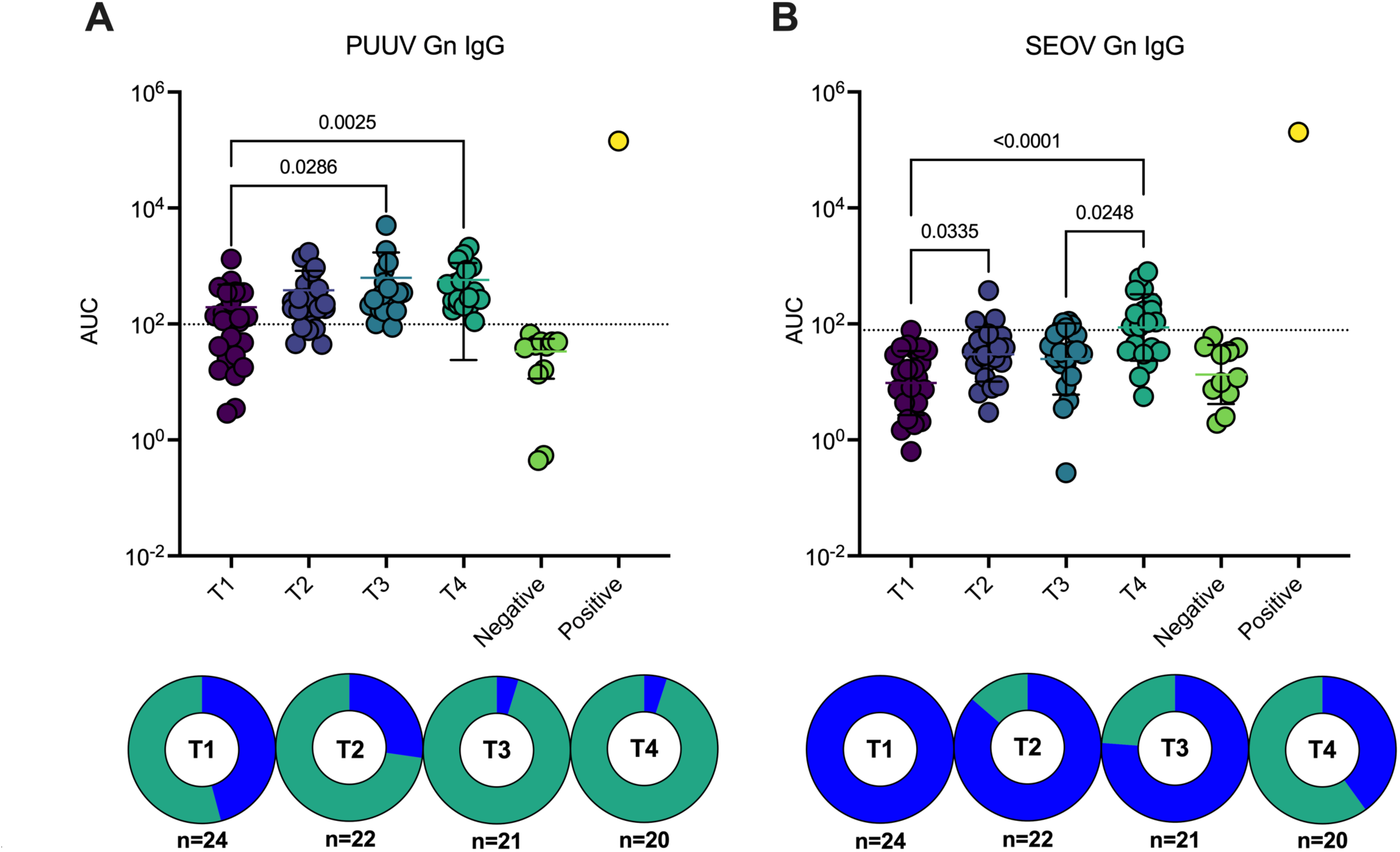
– Patient sera IgG binding to recombinant PUUV and SEOV Gn protein. ELISAs were carried out using patient sera and plates coated with recombinant truncated, Gn of **A)** PUUV and **B)** SEOV. IgG titers are shown as area under the curve (AUC) with the geometric mean and geometric standard deviation denoted by error bars. 11 age and sex matched healthy control sera were included as negative controls and an in-house, anti-PUUV monoclonal antibody was used as a positive control. AUC values were log10 transformed and statistical significance was interrogated via ordinary one-way ANOVA using a Tukey’s multiple comparisons test. Only comparisons with p ≤ 0.05 are shown. Limits of detection (LOD) were calculated for each antigen using the average + 3x standard deviations of the AUC of the negative control sera and are shown as dotted lines on each graph. The percentage of serum samples with LODs above the limit of detection are shown as pie charts, with the percentage of positive samples displayed in green. The n for each time point is displayed below each pie chart.

**Supplementary Figure S2.**
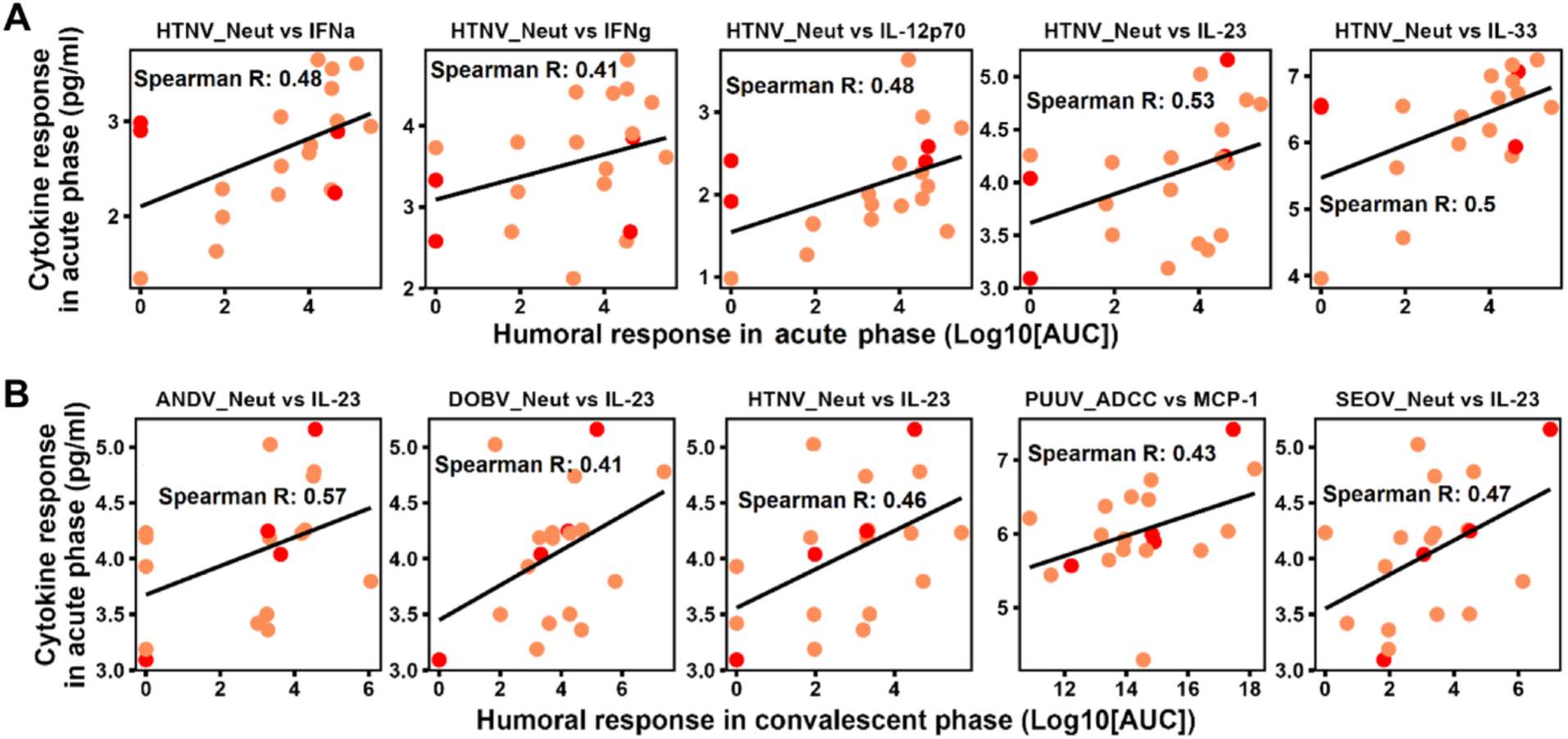
– Correlation analysis between cytokine and humoral responses in acute vs convalescent disease phases. Plots show variables with the highest correlation coefficients between the cytokine concentrations in the acute phase of PUUV infection and the humoral immune response parameters in the **A)** acute (T2) or **B)** convalescent (T4) phases of the infection.

**Table S1.**
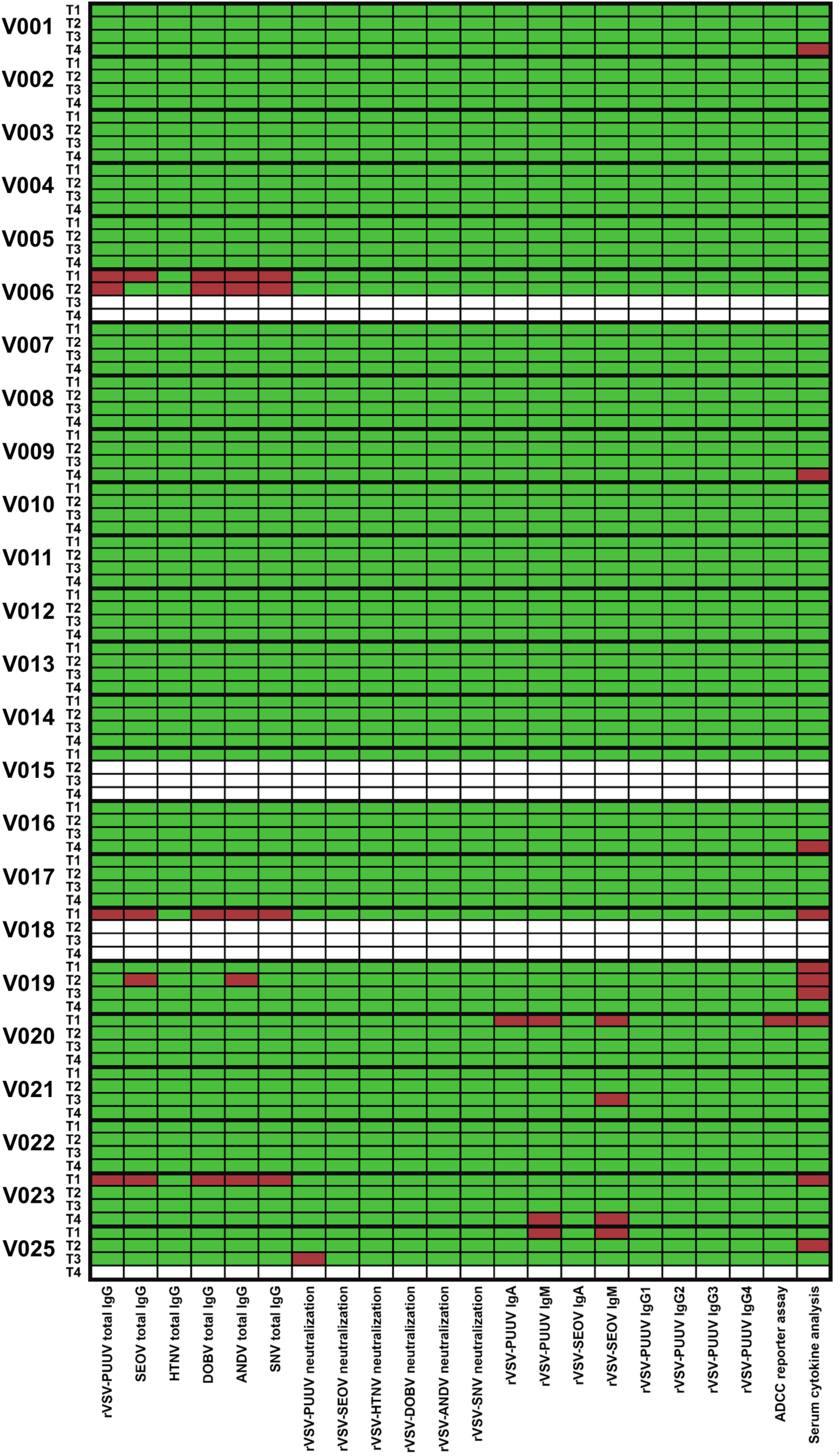
– Patient sera samples that were utilized in this study. Individual patients are shown with the sera samples taken at each of the four time points on the y-axis, while each experimental procedure is shown on the x-axis. Green squares denote where a sample was included, and red squares denote where a sample was not included due to serum scarcity.

